# Dietary and metabolic factors contributing to Barrett’s esophagus: a univariate and multivariate Mendelian randomization study

**DOI:** 10.1101/2023.03.24.23287678

**Authors:** Zijie Li, Weitao Zhuang, Junhan Wu, Haijie Xu, Yong Tang, Guibin Qiao

## Abstract

**Background:** Dietary and metabolic factors have been associated with the risk of Barrett’s esophagus (BE) in observational epidemiological studies. However, the aforementioned associations may be influenced by confounding bias. The present study aimed to evaluate these causal relationships through univariate and multivariate Mendelian randomization (MR) analysis.

**Methods:** Genetic instruments associated with dietary and metabolic factors were obtained in the large-scale genome-wide association studies (GWAS), respectively. Summary data for BE were available from a GWAS of 13,358 cases and 43,071 controls of European descent. Univariable MR analysis was initially performed to estimate the causal relationship between exposures and BE. The inverse-variance weighted (IVW) method was adopted as the primary MR analysis. Multivariate MR analysis was further conducted to evaluate the independent effects of exposures on BE.

**Results:** In univariate MR analysis, BE was causally associated with higher body mass index (odds ratio (OR) = 2.575, 95% confidence interval (CI): 2.301-2.880, P = 7.369E-61), larger waist circumference (OR = 2.028, 95% CI: 1.648-2.496, P = 2.482E-11), and smoking per day (OR = 1.241, 95% CI: 1.085-1.419, P = 0.002). Dried fruit intake showed a protective effect on BE (OR = 0.228, 95% CI: 0.135-0.384, P = 2.783E-08), whereas alcohol drinking, coffee intake, tea intake, fresh fruit intake, and type 2 diabetes mellitus were not associated with BE (P = 0.351, P = 0.458, P = 0.125, P = 0.847, P = 0.413, respectively). No pleiotropy was found in the sensitivity analysis. The relationships of obesity, smoking, and dried fruit intake with BE risk remained strong after adjustment.

**Conclusions:** Our study provided MR evidence supporting obesity and smoking were independent risk factors for BE. Conversely, dried fruit intake was a protective factor for BE.

## Background

Barrett’s esophagus (BE) is characterized by a metaplastic change in the distal esophagus from normal squamous to a specialized columnar epithelium^1^. The prevalence of BE is approximately 1-2% in the population worldwide and is higher in patients with gastroesophageal reflux disease (GERD). BE is a well-known precursor lesion for esophageal adenocarcinoma (EAC), and the process of carcinogenesis has been found to be a sequential progression from metaplasia to dysplasia to cancer^2^. Patients with BE have a 10 to 55-fold increased risk of developing EAC compared to the general population^1^. Therefore, primary prevention of BE is of particular significance.

Numerous observational epidemiological studies have revealed several potential influencing factors for BE, including obesity^3^, type 2 diabetes mellitus (T2DM)^4^, smoking^5^, alcohol drinking^6,7^, coffee and tea intake^8^, and fruit inatke^9^. However, the findings regarding the effects of these dietary and metabolic factors on BE from previous studies are inconsistent even opposite to each other, which make the identification of truly protective and risk factors for BE extremely difficult. For example, previous studies have shown an association between smoking or excessive alcohol drinking and the presence of BE^5,6^, whereas several other case-control studies did not find any relationship between smoking and alcohol drinking and the risk of BE^10,11^. For coffee and tea intake, an Italian study showed that tea intake could reduce the risk of BE while coffee might do the opposite^8^. Nevertheless, another study in the United States did not find any correlation between coffee intake and the risk of BE^12^. In summary, these controversial results have reflected the unignorable limitations of observational epidemiological studies, such as confounding factors and reverse causality, rendering causal inference as a difficult task using classic study methodology^13^.

Since dietary habits and metabolism status are the few potentially modifiable risk factors for BE, it is of great importance to identify their causal relationships using a more robust methodology, in order to achieve a comprehensive understanding and timely intervention for disease prevention. Mendelian randomization (MR) analysis provides an effective alternative analysis method to assess the effect on outcome using single nucleotide polymorphisms (SNPs) as proxies for exposure, reducing the impact of unmeasured confounding and reverse causality^14^. Furthermore, multivariate MR can incorporate genetic variation for multiple exposures into the same model for analysis, rendering it as an effective tool to explore the causal effect of dietary and metabolic factors on broad health-related outcomes^15,16^. The interaction between exposures is eliminated so that the independent effects of individual exposures on the outcome can be estimated simultaneously^17^.

The present study aimed to explore the potential causal relationship between dietary and metabolic factors and the risk of BE through univariate and multivariate MR analysis.

## Method

### Study design

This study was reported according to the strengthening the reporting of observational studies in epidemiology using mendelian randomization (STROBE-MR)^18^, and the study design overview was shown in Figure 1. MR analysis is based on three assumptions: (1) instrumental variables (IVs) are closely associated with exposures; (2) IVs should not be affected by confounders; and (3) IVs only affect BE through exposures^19^. The relationships between exposures and BE were initially explored by univariate MR analysis. Suitable variables were further included in the same model and multivariate MR analysis was utilized to verify the independent effect of each exposure on BE. The present study was based on summary-level data that had been made publicly available, and ethical approval had been obtained in all original studies.

**Figure 1.**
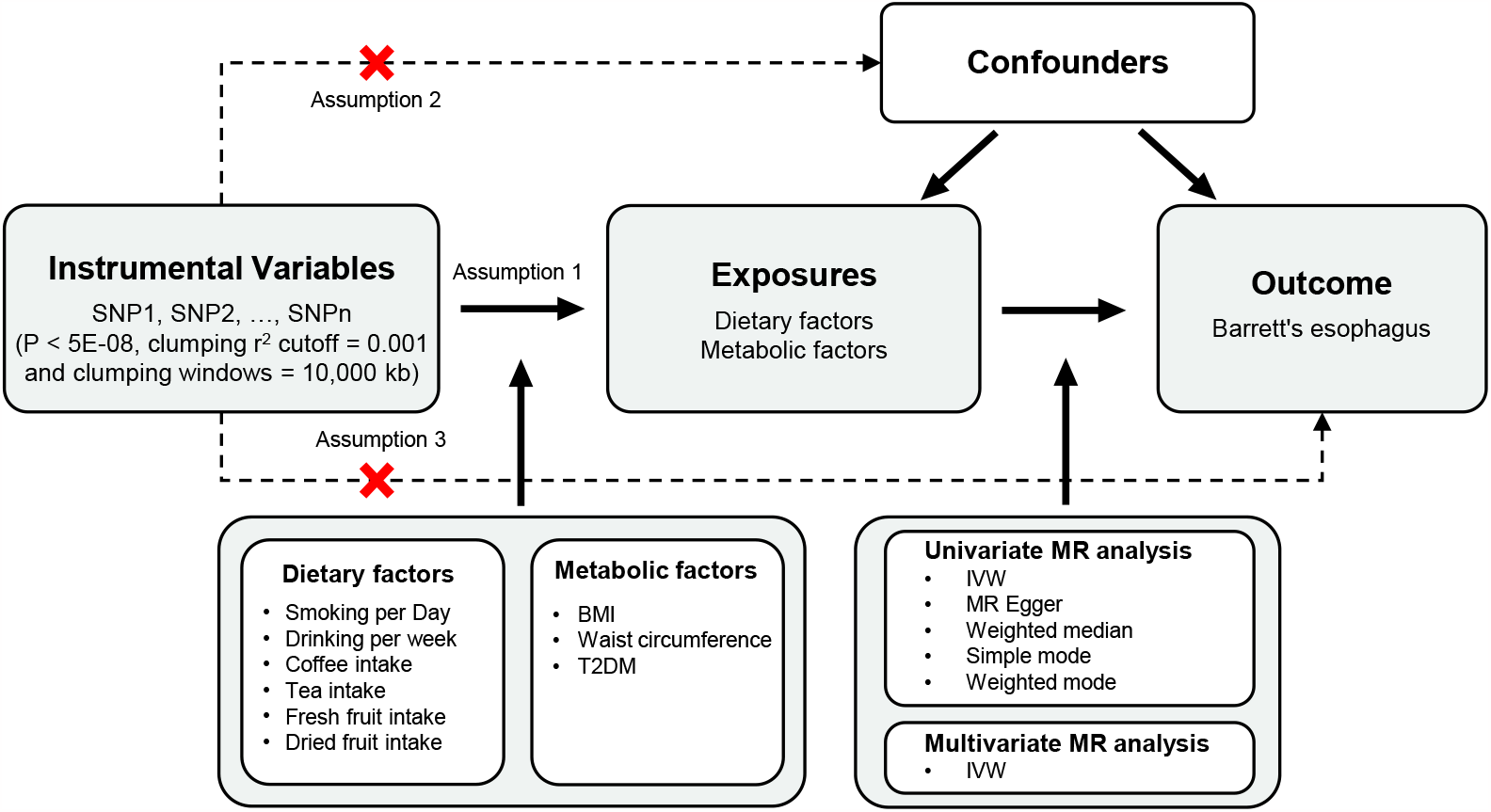
Study design overview. SNPs, single nucleotide polymorphisms; LD, linkage disequilibrium; BMI, body mass index; T2DM, type 2 diabetes mellitus; MR, Mendelian randomization; IVW, inverse variance weighted.

### Data sources

The genetic IVs of exposures were obtained from the summary statistics of published genome-wide association studies (GWAS), which were restricted to European-ancestry individuals to eliminate population stratification bias^20-23^. The data sources of GWAS data were represented in Supplemental Table 1. Summary datasets for body mass index (BMI) and waist circumference were obtained from the Genetic Investigation of ANthropometric Traits (GIANT) consortium^20^. Summary data of T2DM were extracted from the large-scale GWAS including 655,666 participants^21^. Genetic instruments of smoking per day and alcohol drinking per week were extracted from the GWAS and Sequencing Consortium of Alcohol and Nicotine use (GSCAN) consortium^22^. Summary statistics relating to coffee, tea, and fruit were obtained from the Medical Research Council-Integrative Epidemiology Unit (MRC-IEU) consortium. The GWAS summary statistics for BE were obtained from the large-scale published GWAS in the European population, which included 13,358 cases and 43,071 controls^23^.

### Selection of IVs

A series of quality control steps were performed to select eligible SNPs from the GWAS summary data of exposures. All SNPs achieving genome-wide significance (P < 5 × 10^−8^) were screened as IVs. Meanwhile, the linkage disequilibrium (LD) among the SNPs was estimated using 1000 Genomes European panel as the reference population^24^. SNPs in high LD (clumping r^2^ cutoff = 0.001 and clumping windows = 10,000 kb) were excluded to guarantee the independence of IVs. Furthermore, the proxy SNP correlated (r^2^ > 0.8) with the variant of interest was selected when there was no SNP associated with the exposure in the outcome dataset. The exposure SNPs and outcome SNPs were harmonized to maintain concordance of effect alleles, while the palindromic SNPs were further excluded. MR Pleiotropy Residual Sum and Outlier (MR-PRESSO) test was applied to detect and remove outlier SNPs to correct for widespread horizontal pleiotropy and generate estimates without outliers^25^. The R^2^ of each SNP was calculated by the following equation: 2 × Beta^2^× EAF × (1 − EAF), which was used to represent the proportion of variance in an exposure factor explained by the IVs^26^. F-statistics of each SNP were calculated by the following equation: R^2^ × (N – k – 1) ÷ (1 − R^2^). The correlation between the IVs and exposure was considered sufficiently strong when F-statistics > 10, and SNPs with F-statistics < 10 were removed from MR analysis^27^. The final selected SNPs were utilized as the eligible genetic IVs for subsequent MR analysis.

### Statistical Analyses

In this study, the inverse variance weighted (IVW) method was performed as the primary analysis method to estimate the causal relationship between genetic susceptibility to each exposure and the risk of BE^24^. MR Egger, weighted median, simple mode, and weighted mode were considered as complementary methods to infer causality. This causal relationship was considered indicative if the estimates from the IVW method were statistically significant and no conflicting results were found in the other complementary methods. The heterogeneity between SNPs was assessed by Cochran’s Q statistic, and the IVs were deemed to have no heterogeneity when P ≥□0.05^28^. The random-effects IVW model was used to estimate MR effects if significant heterogeneity existed, which was less susceptible to the bias of weaker SNPs-exposure associations^29^. Otherwise, the fixed-effects IVW method was considered as the primary result. The MR-Egger regression intercept was used to assess the horizontal pleiotropy of IVs, with a P value□<□0.05 suggesting pleiotropy^30^. In addition, leave-one-out analysis was performed to assess whether the presence of any outliers would bias the overall MR estimate. The funnel plot and scatter plot were used to visualize the robustness of the results. The asymmetry of the funnel plot indicated the presence of horizontal polymorphism^24^. To further eliminate interaction effects between different exposures, we further performed multivariate MR analysis to adjust for exposures. The Bonferroni method was performed to correct for multiple testing in the study. The association with two-sided P-values < 0.006 (α = 0.05/9) was deemed statistically significant, and P-values between 0.006 and 0.05 were regarded as suggestive evidence of association. Moreover, other statistical tests were two-sided and the statistical significance was set at P-values <□0.05. The odds ratio (OR) was reported per standard deviation increase in the exposure trait. All statistical analyses were conducted using the TwoSampleMR (v0.5.6), Mendelian Randomization (v0.6.0), and MRPRESSO (v1.0) packages in R software (v4.0.0).

The forestploter (v0.1.5) package was employed in drawing forest plot.

## Result

### Univariable MR analysis

After a series of selections of eligible IVs and the exclusion of potentially pleiotropic SNPs, the SNPs closely associated with exposures were applied as IVs (Supplemental Table 2-10). The F-statistic for each SNP was greater than 10 (from 10.2 to 3813.5), indicating all the IVs selected in the MR analysis were of sufficient validity. Univariable MR analysis indicated that the genetic susceptibility of smoking per day, BMI, waist circumference, and dried fruit intake were causally related to the risk of developing BE (Figure 2). The number of smoking per day was causally related to the risk of BE (OR = 1.241, 95% confidence interval (CI): 1.085-1.419, P = 0.002) (Figure 2), and other complementary methods remained in a consistent direction, though not statistically significant (Supplemental Table 11). MR-PRESSO did not detect any influential outliers. In the analysis of the relationship between BMI and BE, the IVW method supported that higher BMI was a risk factor for BE (OR = 2.575, 95% CI: 2.301-2.880, P = 7.369E-61). Likewise, there was a causal relationship between larger waist circumference and the risk of developing BE after the removal of outliers (OR = 2.028, 95% CI: 1.648-2.496, P = 2.482E-11). In the analysis of the relationship between fruit intake and BE, it was worth noting that dried fruit intake was a protective factor for BE (OR = 0.228, 95%CI: 0.135-0.384, P = 2.783E-08). Nevertheless, there was no evidence to support the association between fresh fruit intake and BE (P = 0.847). In addition, alcohol drinking per week, coffee intake, tea intake, and T2DM were not significantly associated with the risk of BE (P = 0.351, P = 0.458, P = 0.125, P = 0.413, respectively). Figure 3 showed the scatter plots of the causal effect estimates of each exposure on the risk of BE, and the forest maps indicated the effect of each SNP for exposures on BE and their whole estimates (Supplemental Figure 1).

**Figure 2.**
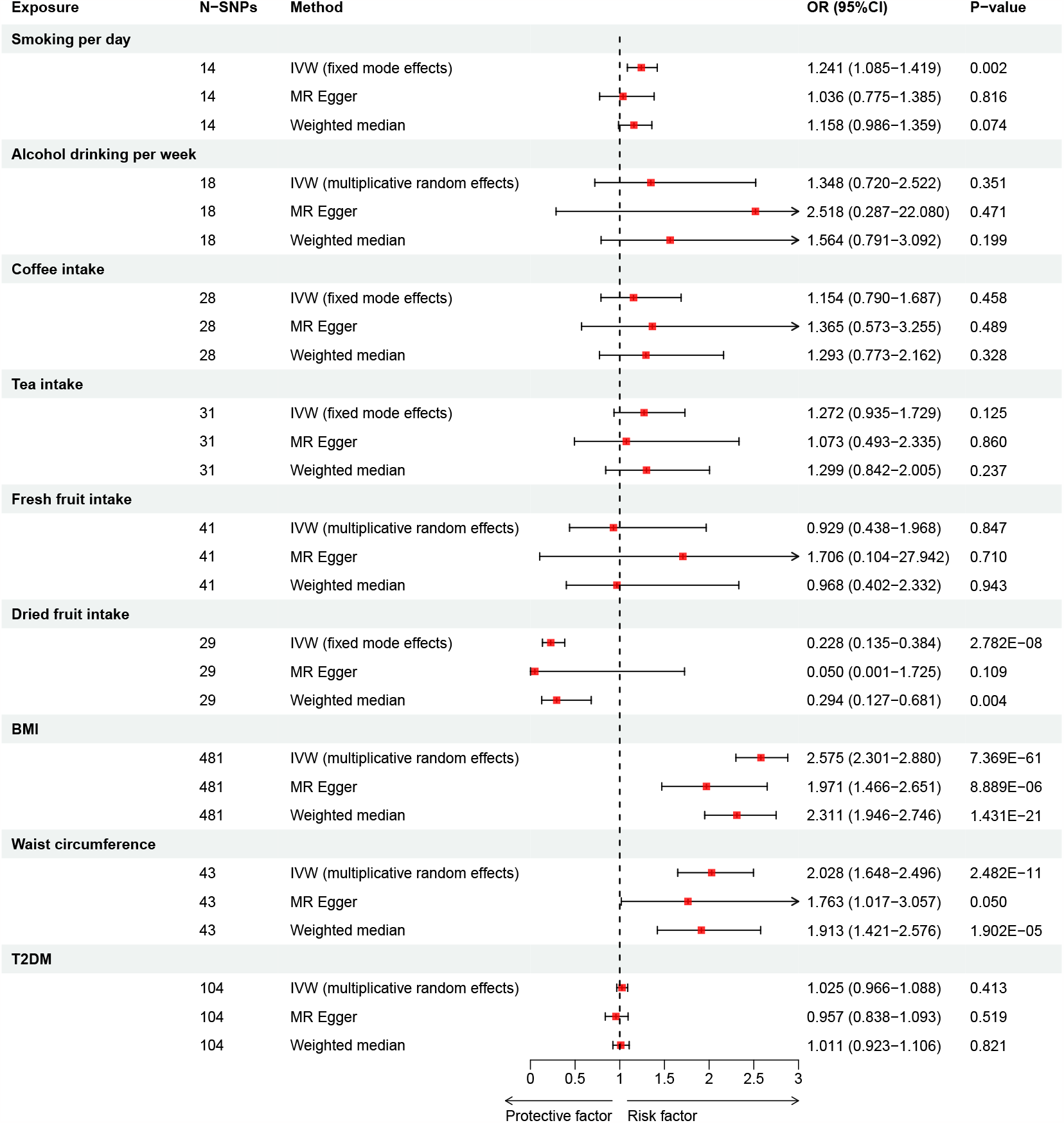
Univariable Mendelian randomization estimated the association between exposures and Barrett’s esophagus. SNPs, Single nucleotide polymorphisms; OR, odds ratio; CI, confidence interval; IVW, inverse variance weighted; BMI, body mass index; BMI, body mass index; T2DM, type 2 diabetes mellitus.

**Figure 3.**
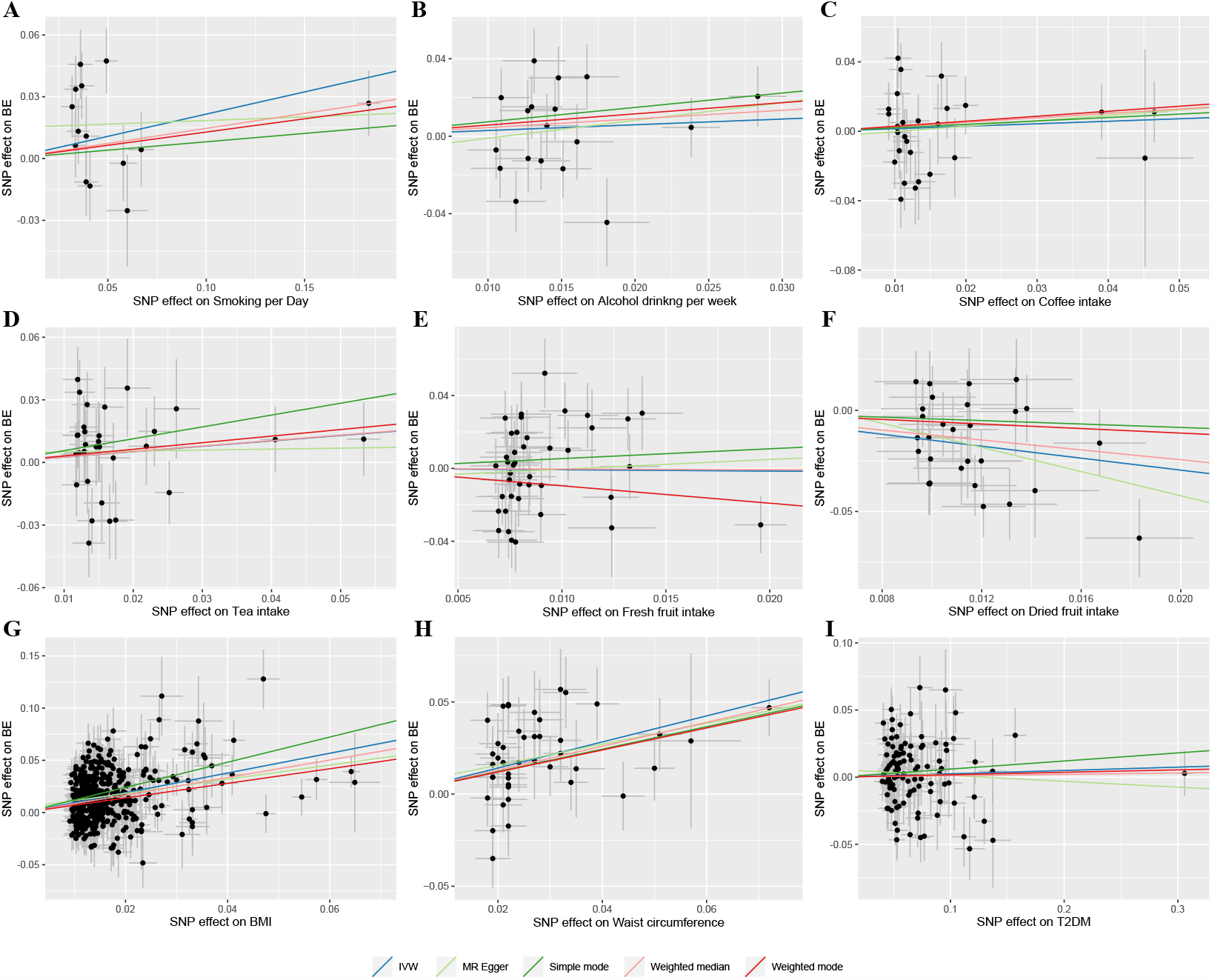
Scatter plots for the association between exposures and Barrett’s esophagus. (A) Smoking per day; (B) Alcohol drinking per week; (C) Coffee intake; (D) Tea intake; (E) Fresh fruit intake; (F) Dried fruit intake; (G) BMI; (H) Waist circumference; (I) T2DM. SNPs, Single nucleotide polymorphisms; BE, Barrett’s esophagus; BMI, body mass index; T2DM, type 2 diabetes mellitus; IVW, inverse variance weighted.

### Sensitivity analysis

The results of the sensitivity analysis were presented in Table 1. There was heterogeneity in the causal estimation of alcohol drinking per week, fresh fruit intake, BMI, waist circumference, and T2DM on BE in Cochran’s Q test (P < 0.05). The MR-Egger intercept analyses showed no evidence of directional pleiotropy in all analyses. The leave-one-out test suggested that the potential relationships between exposures and risk of BE were not driven by any single SNPs (Supplemental Figure 2). The funnel plots showed the symmetrical distribution of points that represented the causal association effect of each SNP, indicating the associations were less affected by potential bias (Supplemental Figure 3).

**Table 1.**
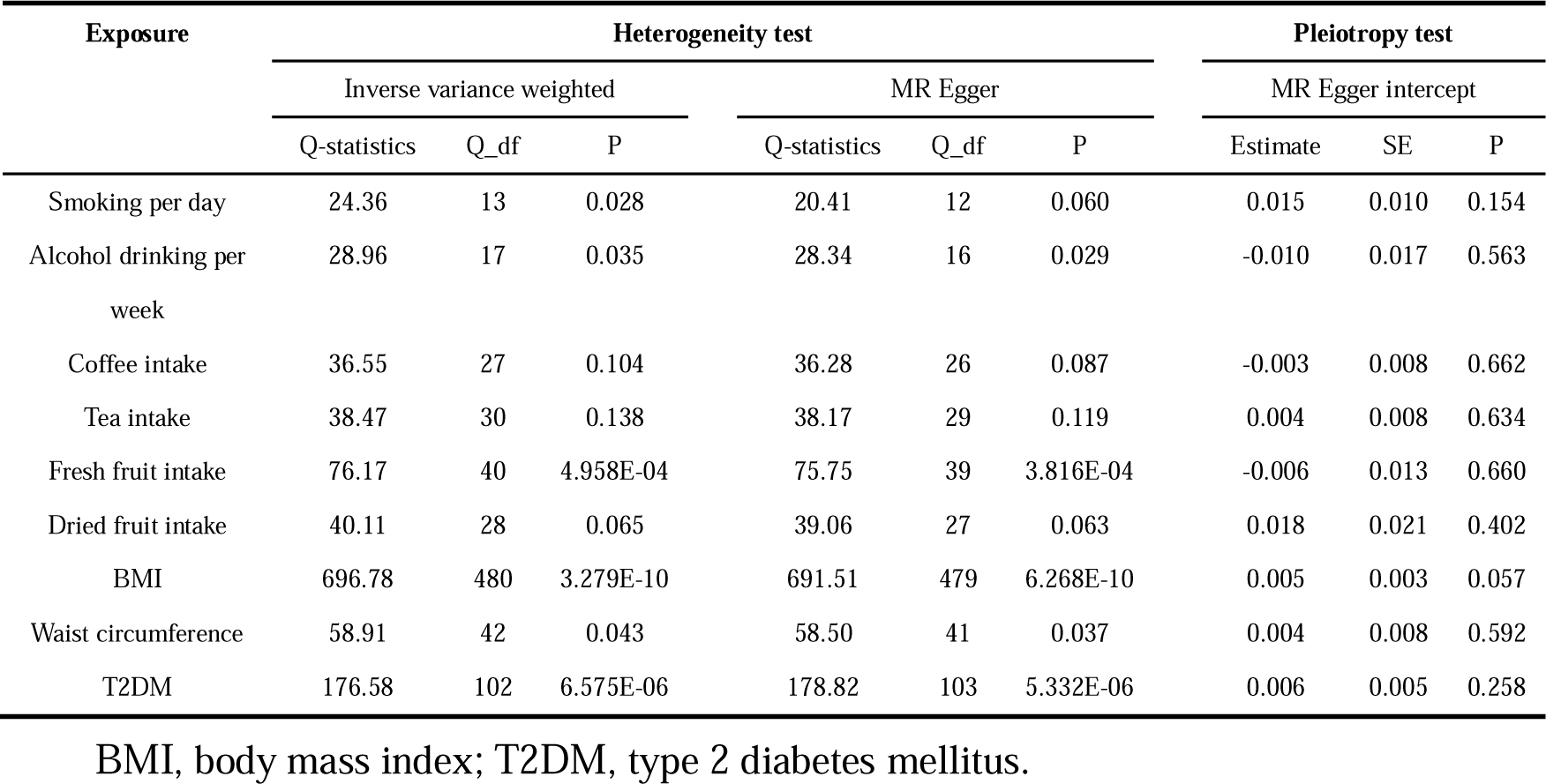
Heterogeneity and horizontal pleiotropy analyses between exposures and risk of Barrett’s esophagus.

| Exposure | Heterogeneity test |  |  |  |  |  | Pleiotropy test |  |  |
| --- | --- | --- | --- | --- | --- | --- | --- | --- | --- |
|  | Inverse variance weighted |  |  | MR Egger |  |  | MR Egger intercept |  |  |
|  | Q-statistics | Q_df | P | Q-statistics | Q_df | P | Estimate | SE | P |
| Smoking per day | 24.36 | 13 | 0.028 | 20.41 | 12 | 0.060 | 0.015 | 0.010 | 0.154 |
| Alcohol drinking per week | 28.96 | 17 | 0.035 | 28.34 | 16 | 0.029 | -0.010 | 0.017 | 0.563 |
| Coffee intake | 36.55 | 27 | 0.104 | 36.28 | 26 | 0.087 | -0.003 | 0.008 | 0.662 |
| Tea intake | 38.47 | 30 | 0.138 | 38.17 | 29 | 0.119 | 0.004 | 0.008 | 0.634 |
| Fresh fruit intake | 76.17 | 40 | 4.958E-04 | 75.75 | 39 | 3.816E-04 | -0.006 | 0.013 | 0.660 |
| Dried fruit intake | 40.11 | 28 | 0.065 | 39.06 | 27 | 0.063 | 0.018 | 0.021 | 0.402 |
| BMI | 696.78 | 480 | 3.279E-10 | 691.51 | 479 | 6.268E-10 | 0.005 | 0.003 | 0.057 |
| Waist circumference | 58.91 | 42 | 0.043 | 58.50 | 41 | 0.037 | 0.004 | 0.008 | 0.592 |
| T2DM | 176.58 | 102 | 6.575E-06 | 178.82 | 103 | 5.332E-06 | 0.006 | 0.005 | 0.258 |

BMI, body mass index; T2DM, type 2 diabetes mellitus.

### Multivariable MR analysis

After adjusting for the potential interactions of variables using multivariate MR analysis, the direct effects of the exposure on BE were explored and the results were shown in Figure 4. In the first multivariate MR model, the causal effect of smoking on BE remained essentially consistent after adjusting for alcohol drinking and BMI (OR = 1.239, 95%: 1.070-1.435, P = 0.004). The effect of obesity on BE was slightly reduced after adjusting for smoking and alcohol drinking (OR = 2.452, 95% CI: 2.154-2.792, P = 8.918E-42). There remained no significant association between alcohol drinking and BE (P = 0.541). In the second multivariate MR model, the protective effect of dried fruit intake on BE was slightly reduced after adjusting for obesity and fresh fruit intake (OR = 0.355, 95% CI: 0.194-0.649, P = 7.742E-04).

**Figure 4.**
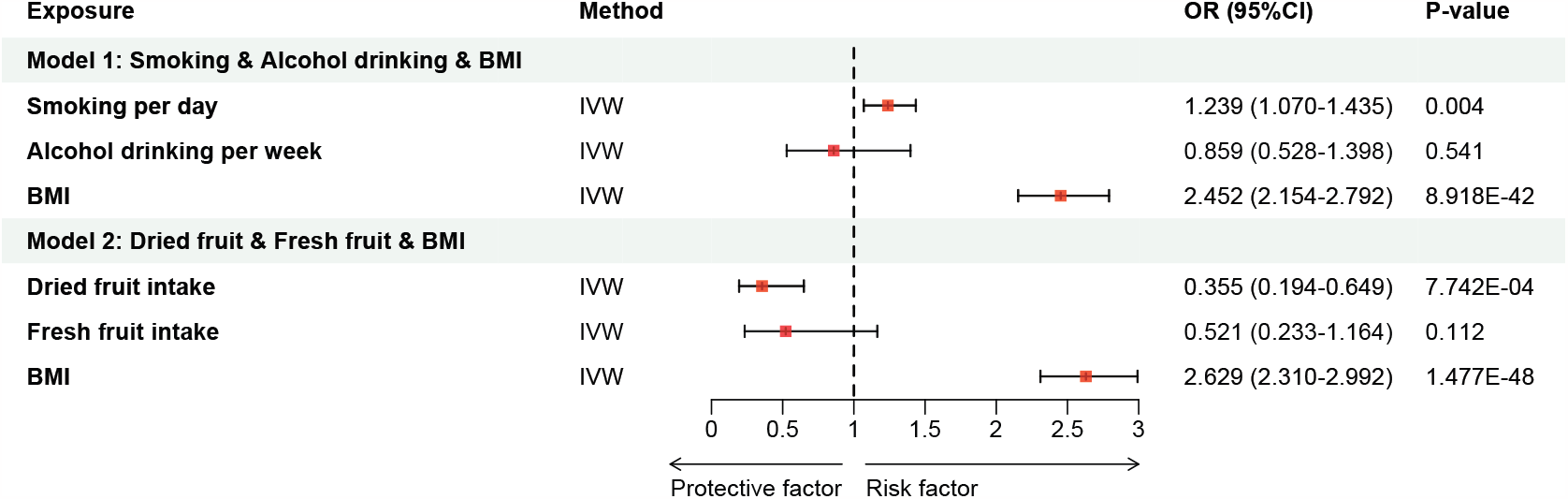
Multivariable Mendelian randomization estimated the association between exposures and Barrett’s esophagus. SNPs, Single nucleotide polymorphisms; OR, odds ratio; CI, confidence interval; BMI, body mass index; IVW, inverse variance weighted.

## Discussion

Compared to previous observational epidemiological studies, MR studies provide genetic proof of potential causality, which avoid the effects of confounding bias and reverse causation. The present MR study supported that the independent causal associations of genetic predisposition to smoking, higher BMI, and larger waist circumference with the risk of developing BE. Remarkably, dried fruit intake played a protective role in BE. However, the MR results demonstrated that alcohol drinking per week, coffee intake, tea intake, fresh fruit intake, and T2DM were not causally related to BE.

Smoking has been consistently shown to be an independent risk factor for BE in various observational studies, which supported our MR finding. A large-scale study covering data from five case-control studies found that smokers had 1.7 times the risk of developing BE compared to never smokers^5^. Another meta-analysis, which included 62 studies with more than 250,157 participants and 22,608 cases, showed that the risk of BE could be effectively reduced by interventions for smoking and the sensitivity analysis for this result was statistically robust^31^. In addition, there was also evidence that smoking primarily contributed to the progression of BE to EAC rather than increasing the risk of Barrett’s esophagus itself^1,32^. Nevertheless, the exact mechanism of smoking on BE remains unclear. Previous studies have suggested that it could be a combination of multiple impacts^33^, including esophageal exposure to N-nitrosamines and genotoxic effects of myosmine^34^, persistent inflammatory irritation of smoking that promoted cell proliferation^35^, as well as the impaired defense mechanism of the esophagus which resulted in GERD^36^.

Our MR study was consistent with the findings of previous observational studies that obesity was an independent risk factor for BE. A meta-analysis including 119,273 subjects found that abdominal obesity was significantly associated with the risk of BE^37^, which mutually confirmed the causal relationship between larger waist circumference and the risk of BE in the current study. Another case-control study using abdominal CT further elucidated that visceral abdominal fat was more strongly associated with BE rather than subcutaneous fat^3^. Several potential mechanisms have been proposed to explain the association, and central obesity may impact BE through both mechanical and metabolic effects^38^. On one hand, central obesity may mechanically disrupt the gastroesophageal junction reflux barrier, leading to an increase in gastroesophageal reflux and hence the development of BE. On the other hand, the visceral abdominal fat compartment mediates the metabolic effect, which releases adipokines and leads to a systemic inflammatory state. Notably, the sleeve gastrectomy in obese patients could alter the anatomy of the His angle, divide the gastric sling fibers and create a narrow stomach in a sleeve fashion, which increases intra-gastric pressure and possibly leads to the aggravation of GERD and BE^39^.

In terms of fruit intake, numerous previous studies have shown that fruit intake can reduce the risk of BE^9,40^. Notably, our study supported that dried fruit was a protective factor for BE, rather than fresh fruit, which might be a novel finding. For fresh fruit, it is widely believed that the intake of fresh fruit can reduce the risk of cancer^41^. Besides, a multicenter case-control study involving 1285 individuals also suggested that the decrease in BE risk was associated with a higher frequency of fresh fruit intake^9^. However, there was still no causal relationship observed between fresh fruit intake and BE in univariate and multivariate MR analysis, which might require larger MR studies as well as better-designed prospective studies to further validate the issue. For dried fruit, traditional dried fruit is formed from fruits such as prunes and grapes by removing water, which is a great source of many micronutrients^42^. Nevertheless, few studies have focused on the association between dried fruits and the risk of BE. Previous studies have suggested that dried fruit had a protective effect on various health-related outcomes such as cardiovascular disease and gastrointestinal disorders^42,43^. A previous MR study has also demonstrated that dried fruit reduced the risk of various cancers such as lung cancer and pancreatic cancer, suggesting a beneficial effect of dried fruit in disease prevention^44^. However, the potential mechanism of the effect of dried fruit on BE needs to be further explored in subsequent studies.

In respect to alcohol drinking and tea intake, most previous studies suggested that there was no association between alcohol or tea drinking and BE^10,12^, which was consistent with the results of the current MR study. However, a few studies have indicated that both wine drinking and tea intake could reduce the risk of BE^8,45^. The possible explanations for the inconsistent results of these observational and the current MR study are as follows: for alcohol drinking, on the one hand, the epidemiological studies may be biased by confounding factors, such as different lifestyles and habits between drinkers and non-drinkers. On the other hand, wine is different from other alcoholic beverages in that moderate consumption of wine might provide a protective effect^45^, while the current MR study did not subdivide the types of alcohol consumed, which might be related to the uncorrelated results. For tea intake, epidemiological studies are likewise biased by confounding factors. Green tea is rich in polyphenols, and catechins are specific polyphenolic compounds that exert anticancer effects by improving the redox state, inhibiting inflammation, and modulating immunity^46,47^. However, summary statistics of tea intake used in the MR analysis included both green and other categories, which could present uncorrelated results. Therefore, further studies are necessary to determine the causal effects of different subtypes of alcohol and tea on BE.

Our MR study found no causal relationship between coffee and BE, while the findings of observational studies regarding the effect of coffee on BE were various. Most studies have concluded that coffee was not associated with BE or had a hazardous effect on BE^8,12^. However, these observational studies are susceptible to confounding factors, and the effect mechanism of various coffee components on BE has not been validated by relevant studies. Notably, dewaxed coffee showed easier digestion and better tolerance, and provided relief for GERD in a recent randomized pilot study^48^, which might further mitigate the risk of developing BE. Coffee as a common beverage has been shown to play an important role in health-related outcomes such as chronic disease and cancer^49^. For example, chlorogenic acid, the main phenolic component of coffee, is a powerful antioxidant and anti-inflammatory agent that plays a key role in reducing the risk of many gastrointestinal diseases and cancers^50^. Therefore, it is highly necessary to identify the specific effects and mechanisms of coffee components on BE.

MR evidence did not support the causal relationship between T2DM and the risk of BE, though observational studies have reported T2DM was a risk factor for BE^4^. Previous studies found that insulin and insulin growth factor 1 were upregulated in BE, which might mediate the development of BE^51^. However, larger sample MR studies as well as experimental studies are needed to validate it in the future.

Our study had several strengths. It was the first application of the univariate and multivariate MR analysis to estimate the causal relationship between various potentially modifiable exposures and BE risk, which minimized potential confounding and reverse causality. The inclusion of summary data based on individuals of European ancestry greatly mitigated the effect of population stratification.

There were several limitations to the present study. First, the specific subtypes of partial exposures remained to be further explored, such as different types of alcohol drinking and tea, and different types of coffee processing affecting its components, which might have different effects on BE. In addition, there were significant heterogeneities in genetic instruments for BMI and T2DM, etc., which might be attributed to the number of participants and SNPs. Given that the included datasets were from participants of European ancestry, it might limit the generalization of the conclusions to non-European populations. The conclusions will be more valid by including data from a larger sample of various ethnicities.

## Conclusion

In summary, our findings provided genetic support that smoking, obesity, and larger waist circumference were risk factors for BE, while dried fruit intake was a protective factor for BE. There was no evidence to support that alcohol drinking, coffee intake, tea intake, fresh fruit intake, and T2DM were associated with the risk of developing BE.

## Data Availability

All data used in the present study were obtained from publicly available GWAS summary statistics, and the datasets presented in this study can be found in online repositories.

## Acknowledgments

The authors acknowledged Jue-Sheng Ong et al. for their contribution, as well as the effort of the GIANT consortium, GSCAN consortium, and MRC-IEU consortium in providing high-quality GWAS data for researchers.

## Author Contributions

Guibin Qiao and Yong Tang designed and supervised the study; Zijie Li and Weitao Zhuang carried out the statistical analyses; Zijie Li, Weitao Zhuang, Junhan Wu and Haijie Xu conducted the study; Zijie Li, Weitao Zhuang, Junhan Wu and Haijie Xu contributed to writing the manuscript; All authors had read and approved the final version of the manuscript.

## Abbreviations

BE: Barrett’s esophagus
GERD: gastroesophageal reflux disease
EAC: esophageal adenocarcinoma
T2DM: type 2 diabetes mellitus
MR: Mendelian randomization
SNPs: single nucleotide polymorphisms
IVs: instrumental variables
GWAS: genome□wide association study
BMI: body mass index
GIANT: Genetic Investigation of Anthropometric Traits
GSCAN: GWAS and Sequencing Consortium of Alcohol and Nicotine use
MRC-IEU: Medical Research Council-Integrative Epidemiology Unit
LD: linkage disequilibrium
MR-PRESSO: MR Pleiotropy RESidual Sum and Outlier
IVW: inverse variance weighted
OR: odds ratio
CI: confidence interval

## Figure Legend

## Supplemental Material

**Supplemental Table 1**. Basic information on the GWAS data applied in this study.

**Supplemental Table 2**. The genetic instruments of smoking per day used in this study.

**Supplemental Table 3**. The genetic instruments of alcohol drinking per week used in this study.

**Supplemental Table 4**. The genetic instruments of coffee intake used in this study.

**Supplemental Table 5**. The genetic instruments of tea intake used in this study.

**Supplemental Table 6**. The genetic instruments of fresh fruit intake used in this study.

**Supplemental Table 7**. The genetic instruments of dried fruit intake used in this study.

**Supplemental Table 8**. The genetic instruments of body mass index used in this study.

**Supplemental Table 9**. The genetic instruments of waist circumference used in this study.

**Supplemental Table 10**. The genetic instruments of type 2 diabetes mellitus used in this study.

**Supplemental Table 11**. Detail of the results of univariate Mendelian randomization.

**Supplemental Figure 1**. Forest plots for the causal effects of dietary and metabolic factors on Barrett’s esophagus. (A) Smoking per day; (B) Alcohol drinking per week; (C) Coffee intake; (D) Tea intake; (E) Fresh fruit intake; (F) Dried fruit intake; (G) BMI; (H) Waist circumference; (I) T2DM. SNPs, Single nucleotide polymorphisms; BE, Barrett’s esophagus; BMI, body mass index; T2DM, type 2 diabetes mellitus; IVW, inverse variance weighted.

**Supplemental Figure 2**. Leave-one-out test plots for the causal effects of dietary and metabolic factors on Barrett’s esophagus. (A) Smoking per day; (B) Alcohol drinking per week; (C) Coffee intake; (D) Tea intake; (E) Fresh fruit intake; (F) Dried fruit intake; (G) BMI; (H) Waist circumference; (I) T2DM. SNPs, Single nucleotide polymorphisms; BE, Barrett’s esophagus; BMI, body mass index; T2DM, type 2 diabetes mellitus; IVW, inverse variance weighted.

**Supplemental Figure 3**. Funnel plots for individual causal effects of dietary and metabolic factors on Barrett’s esophagus. (A) Smoking per day; (B) Alcohol drinking per week; (C) Coffee intake; (D) Tea intake; (E) Fresh fruit intake; (F) Dried fruit intake; (G) BMI; (H) Waist circumference; (I) T2DM. BE, Barrett’s esophagus; BMI, body mass index; T2DM, type 2 diabetes mellitus.

