## Supplemental Figure for "Dietary and metabolic factors contributing to Barrett’s esophagus: a univariate and multivariate Mendelian randomization study"

**Supplemental Figure 1.** Forest plots for the causal effects of dietary and metabolic factors on Barrett's esophagus. (A) Smoking per day; (B) Alcohol drinking per week; (C) Coffee intake; (D) Tea intake; (E) Fresh fruit intake; (F) Dried fruit intake; (G) BMI; (H) Waist circumference; (I) T2DM. SNPs, Single nucleotide polymorphisms; BE, Barrett's esophagus; BMI, body mass index; T2DM, type 2 diabetes mellitus; MR, Mendelian randomization.

**Supplemental Figure 2.** Leave-one-out test plots for the causal effects of dietary and metabolic factors on Barrett's esophagus. (A) Smoking per day; (B) Alcohol drinking per week; (C) Coffee intake; (D) Tea intake; (E) Fresh fruit intake; (F) Dried fruit intake; (G) BMI; (H) Waist circumference; (I) T2DM. SNPs, Single nucleotide polymorphisms; BE, Barrett's esophagus; BMI, body mass index; T2DM, type 2 diabetes mellitus; MR, Mendelian randomization.

**Supplemental Figure 3.** Funnel plots for individual causal effects of dietary and metabolic factors on Barrett's esophagus. (A) Smoking per day; (B) Alcohol drinking per week; (C) Coffee intake; (D) Tea intake; (E) Fresh fruit intake; (F) Dried fruit intake; (G) BMI; (H) Waist circumference; (I) T2DM. BE, Barrett's esophagus; BMI, body mass index; T2DM, type 2 diabetes mellitus.

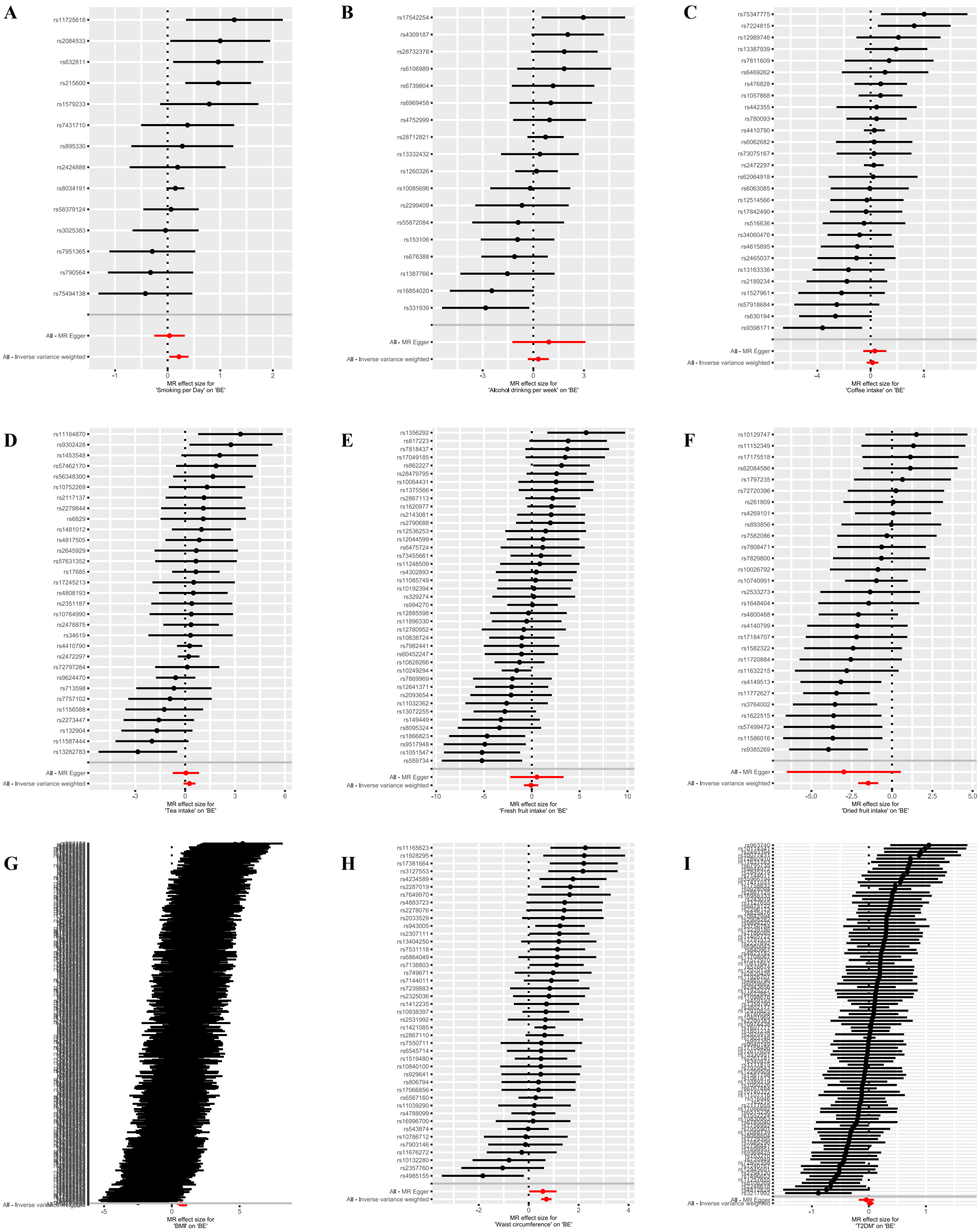


**Supplemental Figure 1.** Forest plots for the causal effects of dietary and metabolic factors on Barrett's esophagus. (A) Smoking per day; (B) Alcohol drinking per week; (C) Coffee intake; (D) Tea intake; (E) Fresh fruit intake; (F) Dried fruit intake; (G) BMI; (H) Waist circumference; (I) T2DM. SNPs, Single nucleotide polymorphisms; BE, Barrett's esophagus; BMI, body mass index; T2DM, type 2 diabetes mellitus; MR, Mendelian randomization.

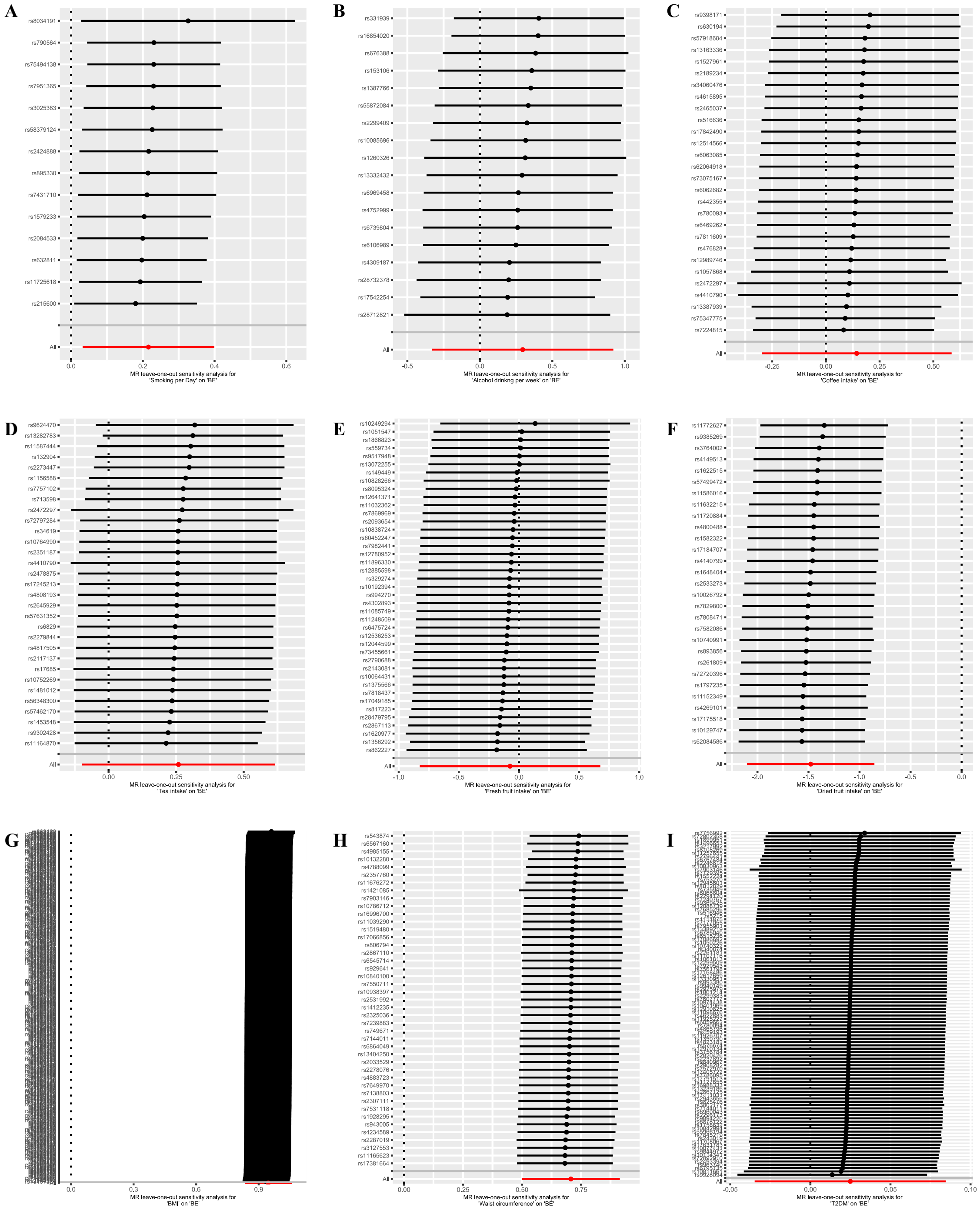


**Supplemental Figure 2.** Leave-one-out test plots for the causal effects of lifestyle and metabolic factors on Barrett's esophagus. (A) Smoking per day; (B) Alcohol drinking per week; (C) Coffee intake; (D) Tea intake; (E) Fresh fruit intake; (F) Dried fruit intake; (G) BMI; (H) Waist circumference; (I) T2DM. SNPs, Single nucleotide polymorphisms; BE, Barrett's esophagus; BMI, body mass index; T2DM, type 2 diabetes mellitus; MR, Mendelian randomization.

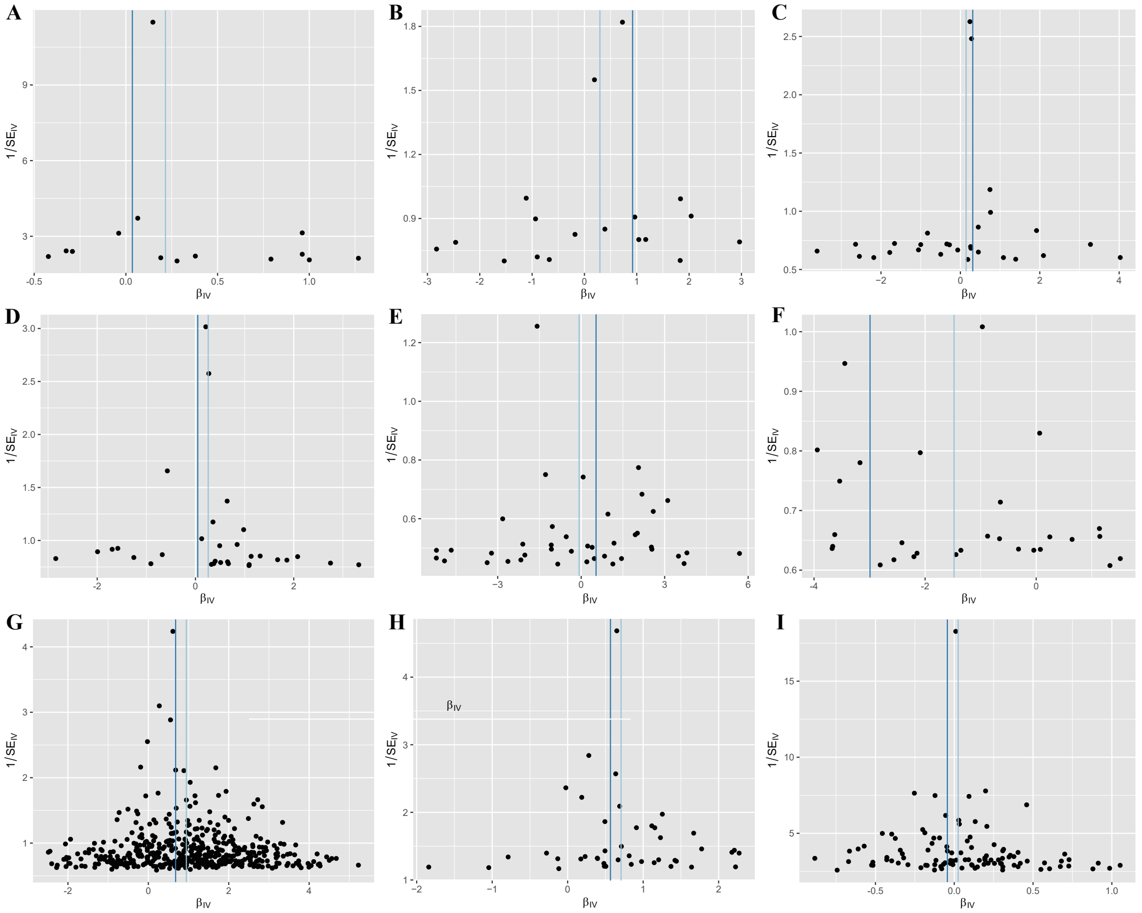


**Supplemental Figure 3.** Funnel plots for individual causal effects of dietary and metabolic factors on Barrett's esophagus. (A) Smoking per day; (B) Alcohol drinking per week; (C) Coffee intake; (D) Tea intake; (E) Fresh fruit intake; (F) Dried fruit intake; (G) BMI; (H) Waist circumference; (I) T2DM. BE, Barrett's esophagus; BMI, body mass index; T2DM, type 2 diabetes mellitus.
